# Genetic variants associated with the risk of stroke in sickle cell disease: a systematic review and meta-analysis

**DOI:** 10.1101/2023.08.11.23294004

**Authors:** Aradhana Kumari, Ganesh Chauhan, Partha Kumar Chaudhuri, Anupa Prasad

## Abstract

**Background:** Sickle cell disease (SCD) is the commonest cause of stroke in children. As it is a rare disease, studies investigating the association with complications like stroke in SCD have small sample sizes. Here, we performed a systematic review and meta-analysis of the studies exploring an association of genetic variants with stroke to get a better indication of their association with stroke.

**Methods:** PubMed and Google Scholar were searched to identify studies that had performed an association analysis of genetic variants for the risk of stroke in SCD patients. After screening of eligible studies, summary statistics of association analysis with stroke and other general information were extracted. Meta-analysis was performed using the fixed effect method on the tool METAL and forest plots were plotted using the R program. The random effect model was performed as a sensitivity analysis for loci where significant heterogeneity was observed.

**Results:** 408 studies were identified using the search term and after screening 39 studies that cumulatively analysed 11,780 SCD patients were included. These 39 studies included a total of 2,401 SCD patients with stroke, predominantly included individuals of African ancestry (N=16). Three of these studies performed whole exome sequencing while 36 performed single nucleotide-based genotyping. Though the studies reported association with 109 loci, meta-analyses could be performed only for 12 loci that had data from two or more studies. After meta-analysis we observed that four loci were significantly associated with risk for stroke: -α3.7kb *Alpha-thalassemia deletion* (P= 0.00000027), rs489347-*TEK* (P= 0.00081), rs2238432-*ADCY*9 (P= 0.00085) and rs11853426-*ANXA2* (P= 0.0034).

**Conclusion:** Ethnic representation of regions with a high prevalence of SCD like the Mediterranean basin and India needs to be improved for genetic studies on associated complications like stroke. Larger genome-wide collaborative studies on SCD and associated complications including stroke needs to be performed.

## 1. Introduction

The global burden of sickle cell disease (SCD) is expected to cross 400,000 by 2050, which will be a major burden on the health system (1,2). SCD is prevalent mainly in ethnic populations of African, Hispanic, Southeast Asian, Indian, and Mediterranean descent(3). It is associated with multiple vascular complications like acute chest syndrome, painful crisis, priapism, avascular necrosis, and stroke (4). Stroke affects 11% of SCD patients before they attend adulthood (5) and is a major contributor to the health burden in SCD(3). Though SCD is caused by the replacement of glutamate residue (Glu6) by valine (Val6) due to a point mutation in the HBB gene (6), there may be multiple genetic variants across the genome that modulate disease onset, severity, and associated complications(7). The pathophysiology of stroke in SCD displays a complex interaction between the genes involved in endothelial dysfunction(8,9), haemolytic episodes(10), coagulation pathways(11–13), inflammatory processes(9,14) and intermediary metabolism(15,16). Multiple studies have investigated the association of variants in *TNF-alpha* (14,16–18), *MTHFR* (19–22), and *VCAM-1*(17,18,23) with stroke in SCD patients. Similarly, studies have also investigated association of genetic variants and have shown association of *ADCY9*(24)*, TEK*(18,24), *GOLGB1*(15,18)*, ENPP1*(15,17,18)*, PON1*(12,17)*, ANXA2*(12,16)*, KL*(25)*, Alpha 3.7 thal del*(21)*, βS Haplotype*(26)*, SOD2*(27)*, and BCL11*(28) with stroke. This list not only includes single nucleotide polymorphism genotyping studies but also includes genome-wide association studies (29,30), whole exome studies (15,31), and whole genome studies(32). However, most of these studies have been performed on a smaller sample size due to the rare occurrence of stroke in SCD. Meta-analysis of these smaller studies will provide good pieces of evidence about the status of the association of these genetic variants with stroke in SCD. Systematic review and meta-analysis of these various genetic modulators of stroke in SCD patients are lacking. Here we present a comprehensive systematic review and meta-analysis of genetic studies published between 1999 and 2022 which have investigated the association of various genetic variants that modulate the risk of stroke in SCD patients.

## 2. Methods

### 2.1 Search Criteria

The study was performed conforming to Preferred Reporting Items for Systematic Reviews and Meta-analysis (PRISMA) Guidelines 2020. The search engines PubMed (https://pubmed.ncbi.nlm.nih.gov/) and Google Scholar (https://scholar.google.com/) were searched for relevant articles using specific search terms for articles with last date of access on 17^th^ February 2022. The specific search terms used were “((stroke) AND ((sickle cell anemia) OR (sickle cell disease))) AND (genetics)” (**Figure 1**). Important studies resulting from the search of the reference section of articles available through search engines were also included (**Figure 1**). The research protocol of this systematic review and meta-analysis was registered on the PROSPERO (https://www.crd.york.ac.uk/prospero/) web portal with registration number “CRD42022311257”.

**Figure 1.**
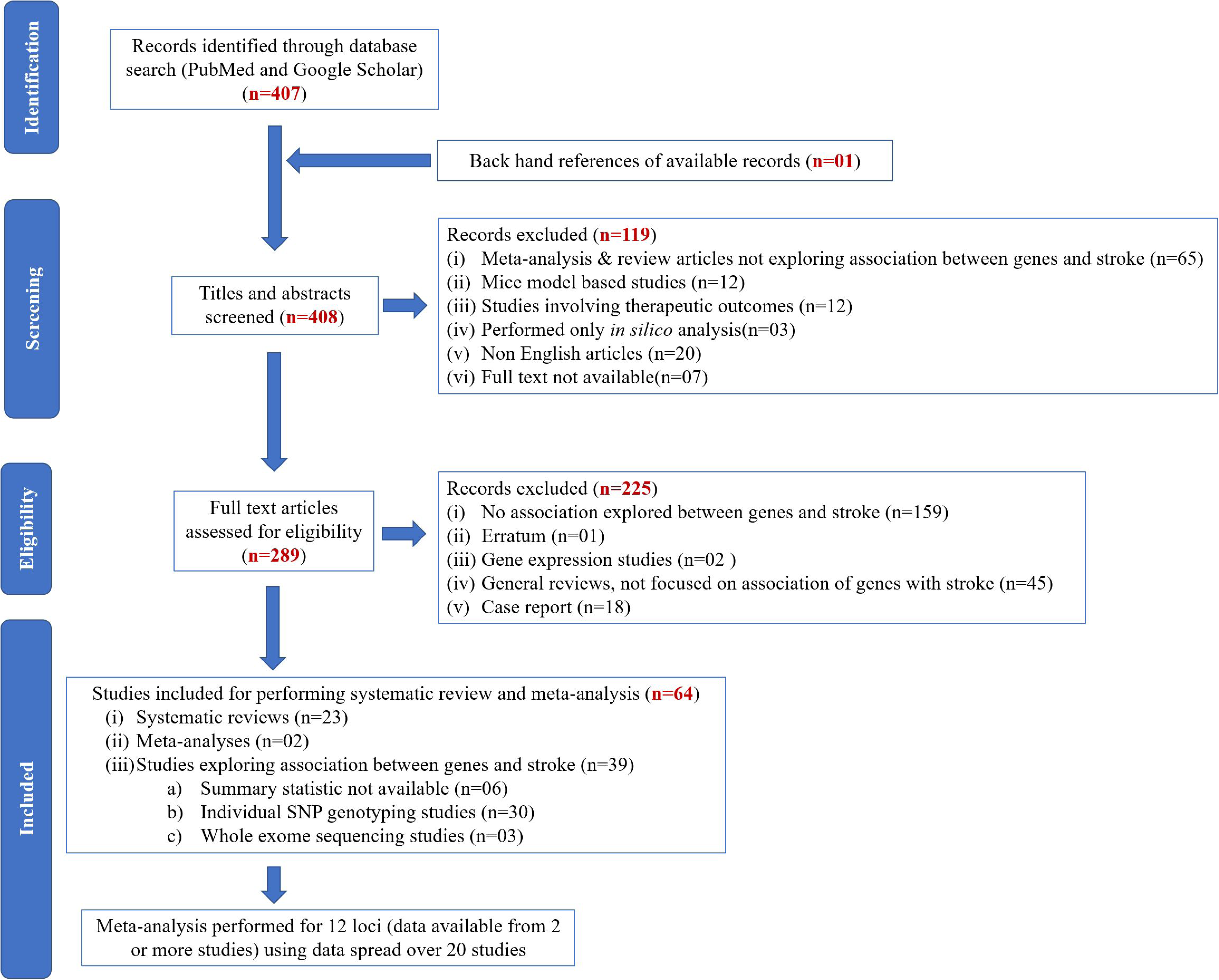
Study Flow diagram representing the study selection and inclusion.

### 2.2 Inclusion and exclusion criteria for selecting studies

The main criteria for selecting studies were that the studies must have performed an association analysis of genetic variants with stroke in a study population of SCD. Any method of availing information on genetic variation be it by genotyping of single or multiple polymorphisms, genome-wide genotyping for GWAS studies, whole exome sequencing studies, or whole genome sequencing studies was included. For all the studies, cases were SCD patients affected with stroke while the control group was SCD patients not affected with stroke. Stroke was defined by WHO criteria(33) or imaging studies performed in individual studies. Stroke subtypes included ischemic and haemorrhagic stroke. The result section, tables, and figures of the articles were screened to extract information on the number of study participants in case and control groups, odds ratio, 95% confidence intervals, beta, standard errors, P-values, and allele frequencies. In certain situations, odds ratio, 95% confidence intervals, beta, and standard errors were calculated using the number of participants with different genotypes in each group of cases and controls. We excluded all case reports, studies reported in languages other than English, studies with no data on association statistics of genetic variants with stroke, and studies where full text was not available. All systematic reviews and meta-analyses were included as part of the search strategy but data for meta-analysis in the current study were included only from independent studies. A detailed list of all studies available through the search engines and the reason for exclusion are presented as **Supplementary Table S1**.

### 2.3 Data Extraction

A data extraction form was created and included information on the journal, lead author, year of publication, ethnicity, study design, total number of participants as cases and controls, males (%), mean/median age, stroke type, genes/variants, ORs, 95%CIs, alleles, frequency of effect allele, effect size in terms of beta of regression and standard error. The race and ethnicity were sourced either from individual study data included in the meta-analysis or based on the literature survey about the study population. Data were extracted by two authors (AT and AP) independently and any ambiguity was settled by consulting a third author (GC).

### 2.4 Quality of studies and risk of bias assessment

Quality of studies and risk of bias assessment were performed using the Q-Genie(35) and modified ROBINS-I tools (36,37). The Q-Genie categorized studies as either good, moderate, or poor quality and the ROBINS-I tool had six domains of assessment and it rated the risk of bias into either low, moderate, serious, or critical. The overall risk of bias for a study was considered severe or critical if any of the six domains in the ROBINS-I tool had severe or critical risk.

### 2.5 Statistical Analysis

Meta-analysis was performed using the fixed effect inverse variance-weighted meta-analysis process using the tool METAL (http://csg.sph.umich.edu/abecasis/metal/) with release date 25^th^ March 2011(34). Beta and standard error obtained from regression analysis or beta and standard error derived from odds ratio and 95% confidence intervals were used for performing a meta-analysis. The heterogeneity of effects between studies was calculated using I^2^ statistics and Cochrane’s Q test P-value as implemented in the tool METAL. For genetic variants which showed high heterogeneity, we also performed a meta-analysis under the random effect model using METAL. Individual forest plots were created for each genetic variant using R version 3.6.3 and used meta-analysis statistics derived from METAL. To check publication bias among the studies included in the meta-analysis, funnel plot was created using RevMan software version 5.4.1.

## 3. Results

### 3.1 Search Results

Using the search term, we retrieved 407 articles and one other article was included based on backhand references search of selected articles, thus a total of 408 studies were evaluated (**Figure 1 and Supplementary Table S1, S2 Excel**). A review of the title and abstract alone led to the exclusion of 119 articles. After screening the full text, another 225 articles got excluded and 64 articles remained that were included for systematic review and meta-analysis performed in the current study (**Figure 1 and Supplementary Table S1, S2 Excel**). Among the 64 articles used in the current study, all included an exploration of genetic variants with stroke in a study population of sickle cell disease patients. These 64 articles included 23 systematic reviews, two meta-analyses, and 39 studies that performed the association of genetic variants with stroke in SCA patients (**Figure 1 and Supplementary Table S2 Excel**). The 39 studies included a total of 11,780 SCA patients, where 2401 patients had suffered from stroke and 9,379 did not suffer from stroke (**Table 1 and Supplementary Table S3 Excel**). The majority of the studies were from Brazil (N=17) and the USA (N=11), the rest from various countries in Africa, one each from the Middle East and Europe, and none from India. Individuals of African ancestry represented the majority of participants in these studies (16 studies and 5,097 participants).

**Table 1:**
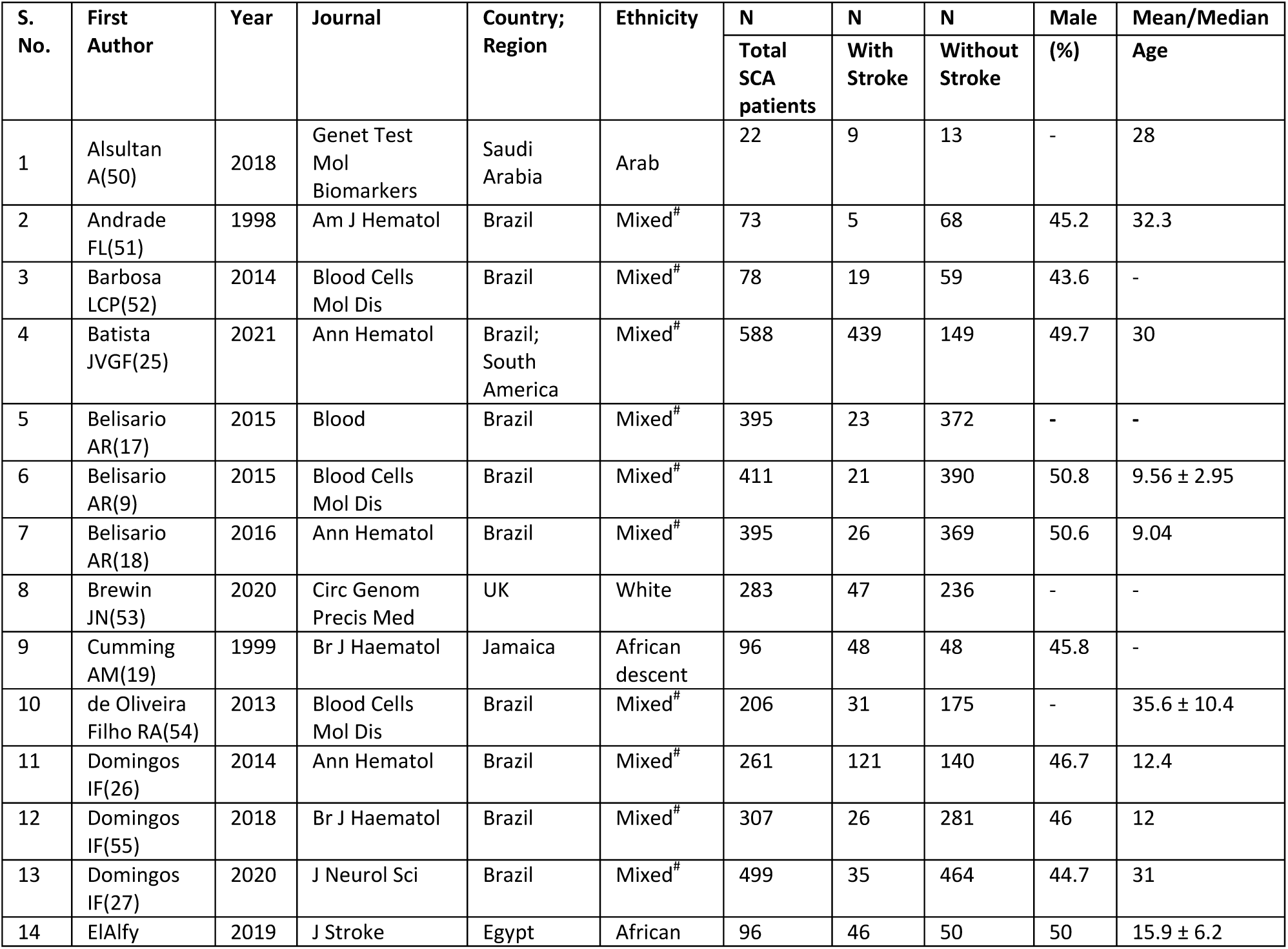
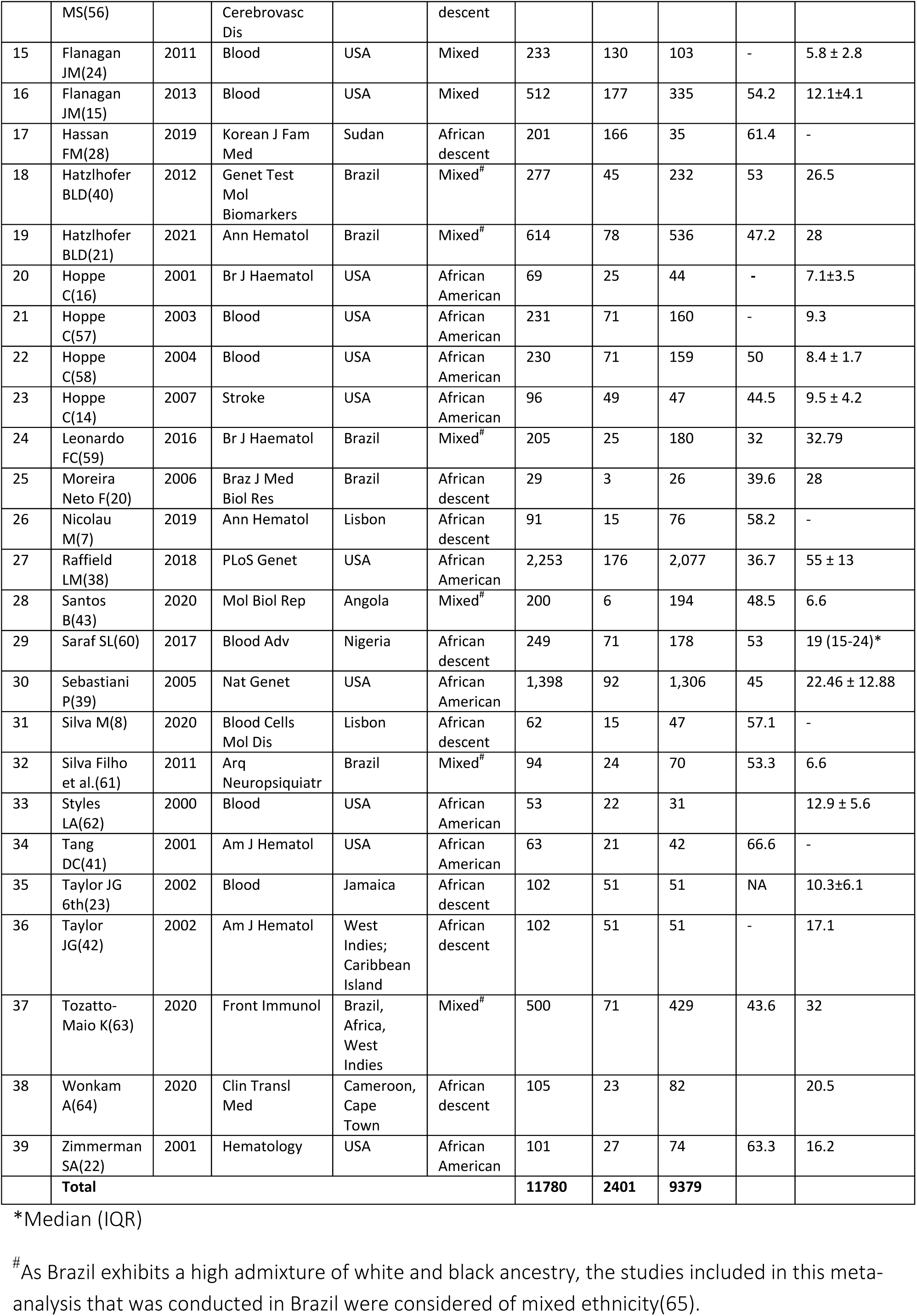
Characteristics of the studies performing association of genetic variants with stroke in SCA patients.

Among the 39 studies that analysed the association of genetic variants with stroke, there were six studies where summary statistics like Odd Ratio, 95% Confidence interval, Beta effect size, and Standard error using the frequentist approach were not available. Thus, data from 33 studies which included 3 whole exome sequencing and 30 individual SNP genotyping were eligible for meta-analysis where they provided summary statistics for association with stroke of one more genetic variant. Upon screening of the 109 genetic variants studied in these 33 studies, we observed that there were only 12 genetic variants where data was available from more than one study and this data was spread across 20 studies **(Figure 1 and Supplementary Table S3 Excel)**. Data were available for 97 other genetic variants which were from only one study; hence meta-analysis could not be performed **(Supplementary Table S4 Excel)**.

### 3.2 Association of genetic variants with Stroke and Meta-analysis

Details of the 109 genetic variants where an association with stroke was performed are provided in **Supplementary Table S4 Excel**. After performing a meta-analysis using a fixed effect model, we observed that seven among the 12 loci were associated with stroke in SCA patients at P<0.05. However, after performing Bonferroni correction for the number of loci tested only four loci remained significant (0.05/12=0.0041) **(Table 2)**. These four loci included -α3.7kb *Alpha-thalassemia deletion* (P=0.0000002668), rs489347-*TEK* (P=0.0008127), rs2238432-*ADCY*9 (P=0.0008553) and rs11853426-*ANXA2* (P=0.003434). No significant heterogeneity was observed for any of the genetic variants. We also performed a random effect meta-analysis as a sensitivity analysis and, the association of the other four variants which did not show significant heterogeneity remained significant in the random effect meta-analysis **(Table 2 and Figures 2a, 2b, 3a, 3b)**. As we observed no heterogeneity in the effects among the studies included in this meta-analysis, we did not employ subgroup analysis.

**Figure 2a.**
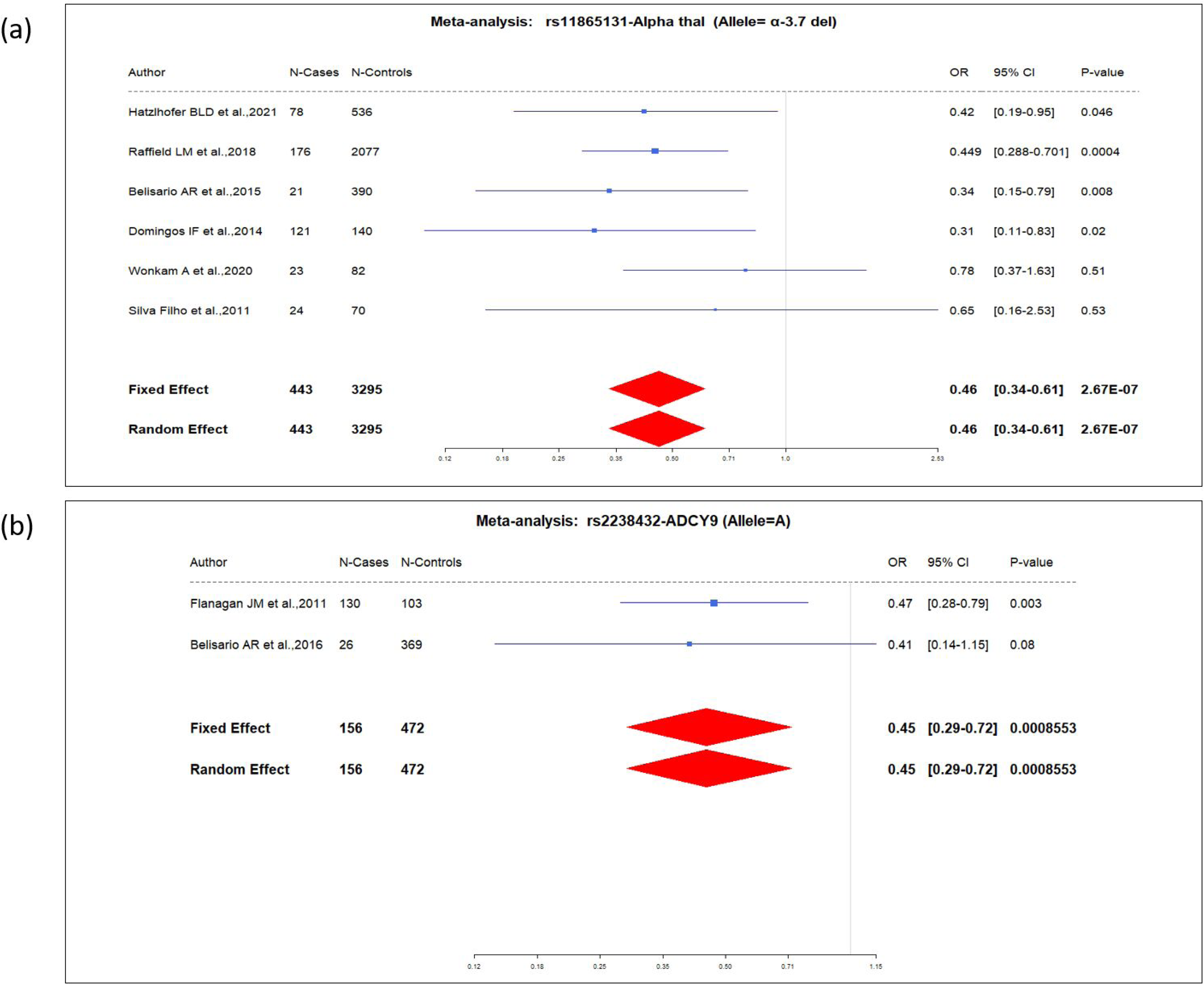
Forest plot showing pooled odds ratio of rs11865131-Alpha thalassemia (α 3.7 kb del) for association with stroke in sickle cell disease.

**Figure 2b.**
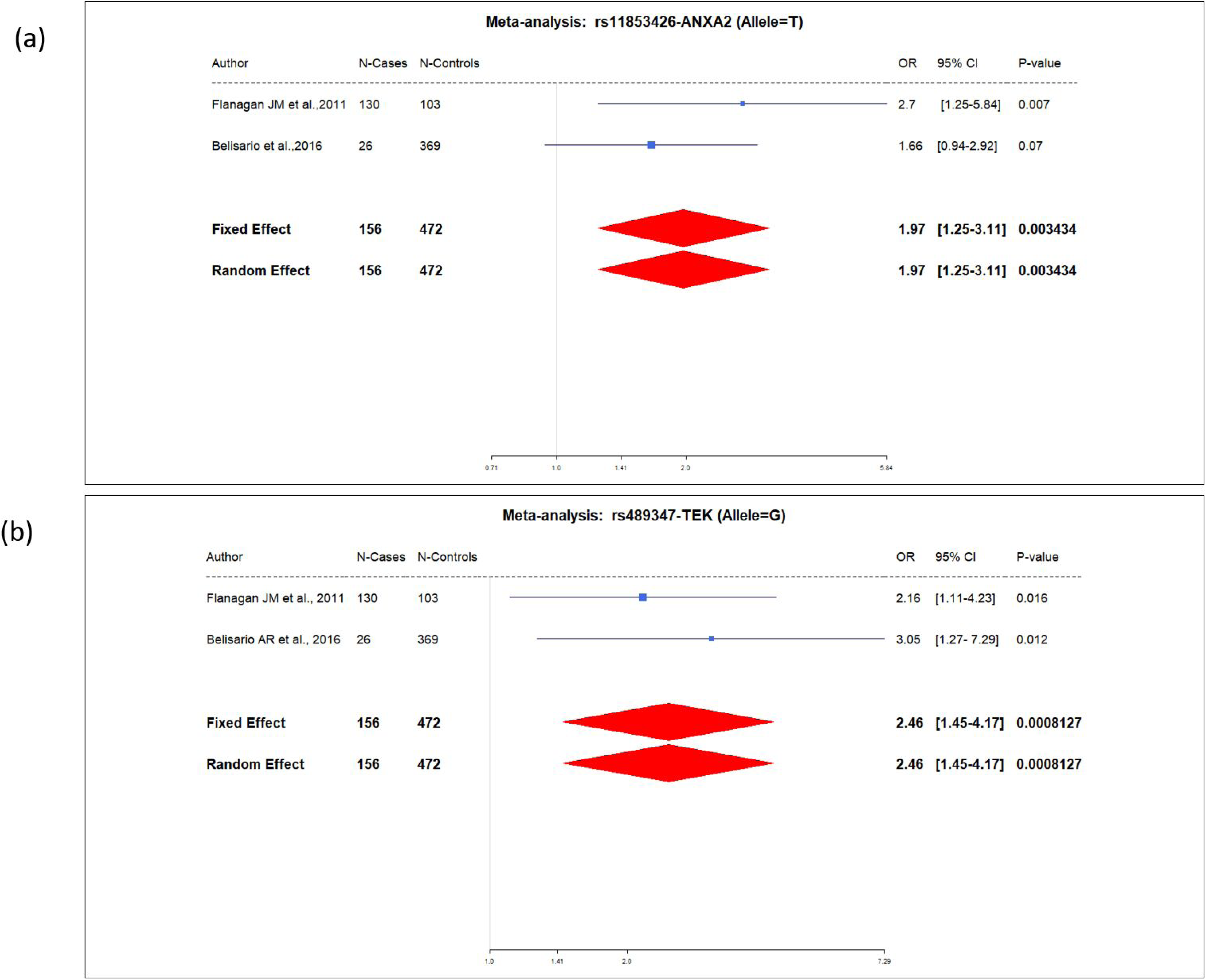
Forest plot showing pooled odds ratio of rs2238432-ADCY9 for the association with stroke in sickle cell disease.

**Figure 3a.** Forest plot showing pooled odds ratio of rs11853426-ANXA2 for association with stroke in sickle cell disease.

**Figure 3b.** Forest plot showing pooled odds ratio of rs489347-TEK for association with stroke in sickle cell disease.

**Table 2:**
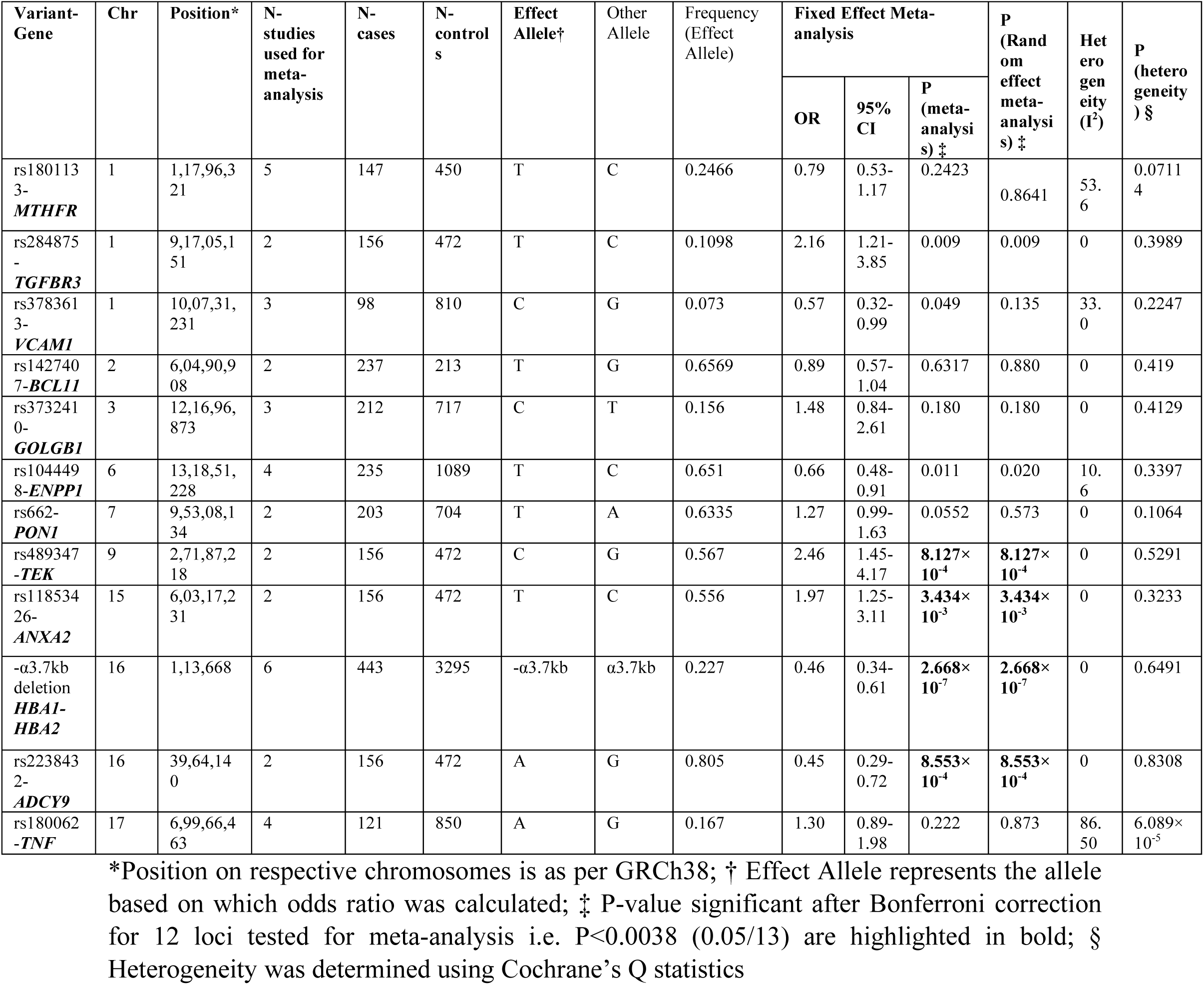
Meta-analysis of results of 12 loci having data from two or more studies.

In this meta-analysis, we did not observe a significant association of the genetic variations in *BCL11A,* ENPP1, GOLGB1, VCAM1, PON1, MTHFR, TNF-α, and TGFBR3 genes with stroke in SCD after the adjustment of statistical significance test for multiple comparisons. Forest plots of the 12 loci are presented in **Supplementary Figures S1 to S12**.

### 3.3 Publication bias

Deeks funnel plot was used to check the publication bias. The symmetric shape of the funnel plot indicated a low probability of publication bias in the studies included in the meta-analysis of the four genetic variants **(Supplementary Figure S13).**

### 3.4 Study quality and risk of bias assessment

Using the Q-Genie tool, four studies were classified as having good quality(7,15,38,39), twenty-eight as moderate quality (8,12–16,18,20,21,30–47,60), and seven as poor quality(17,19,23,28,40–42). The overall risk of bias was rated as low, moderate, severe, or critical. It was predominantly affected by selection bias, confounding bias, and bias due to the assessment of outcome. **(Table 3)**.

**Table 3:**
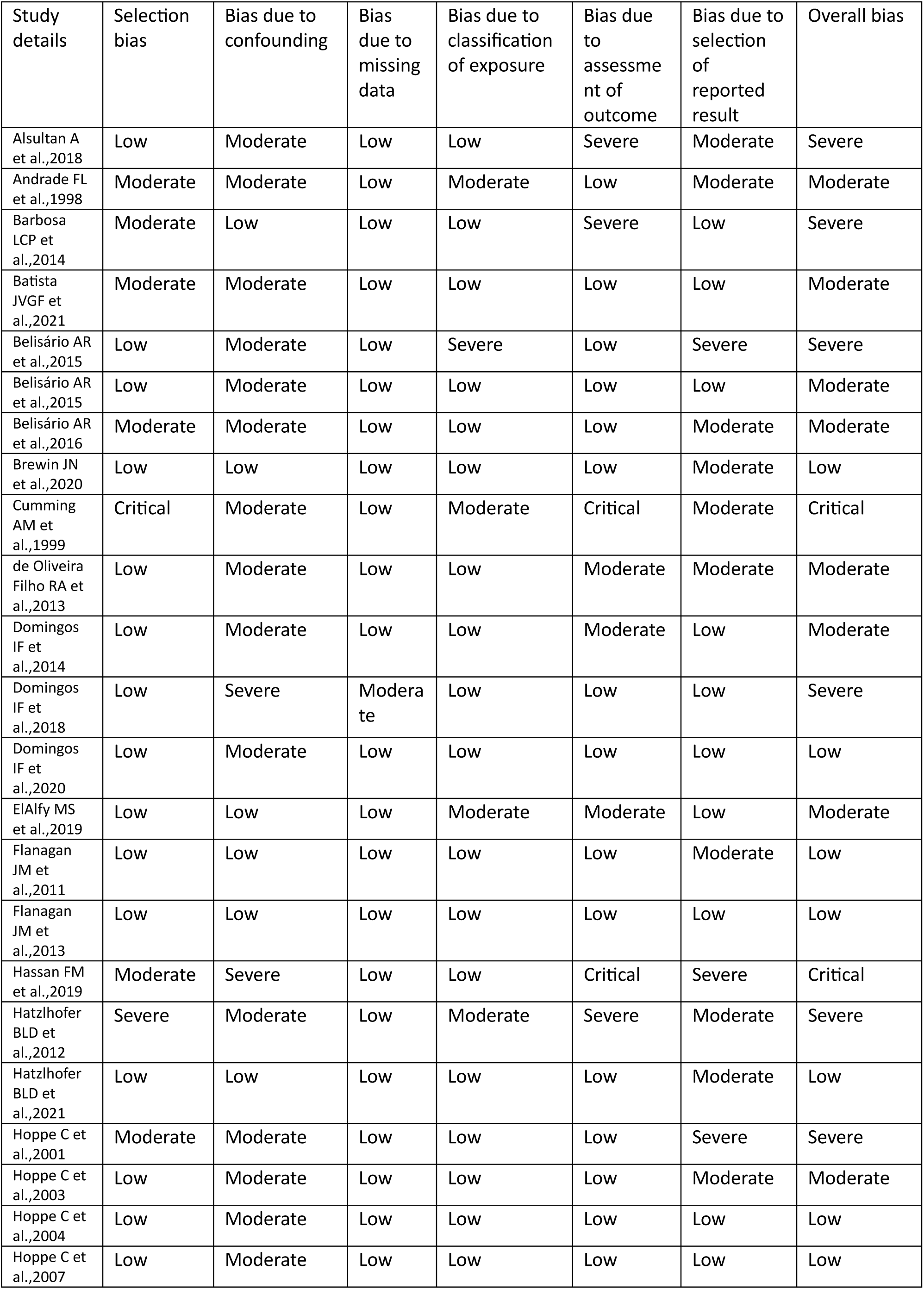
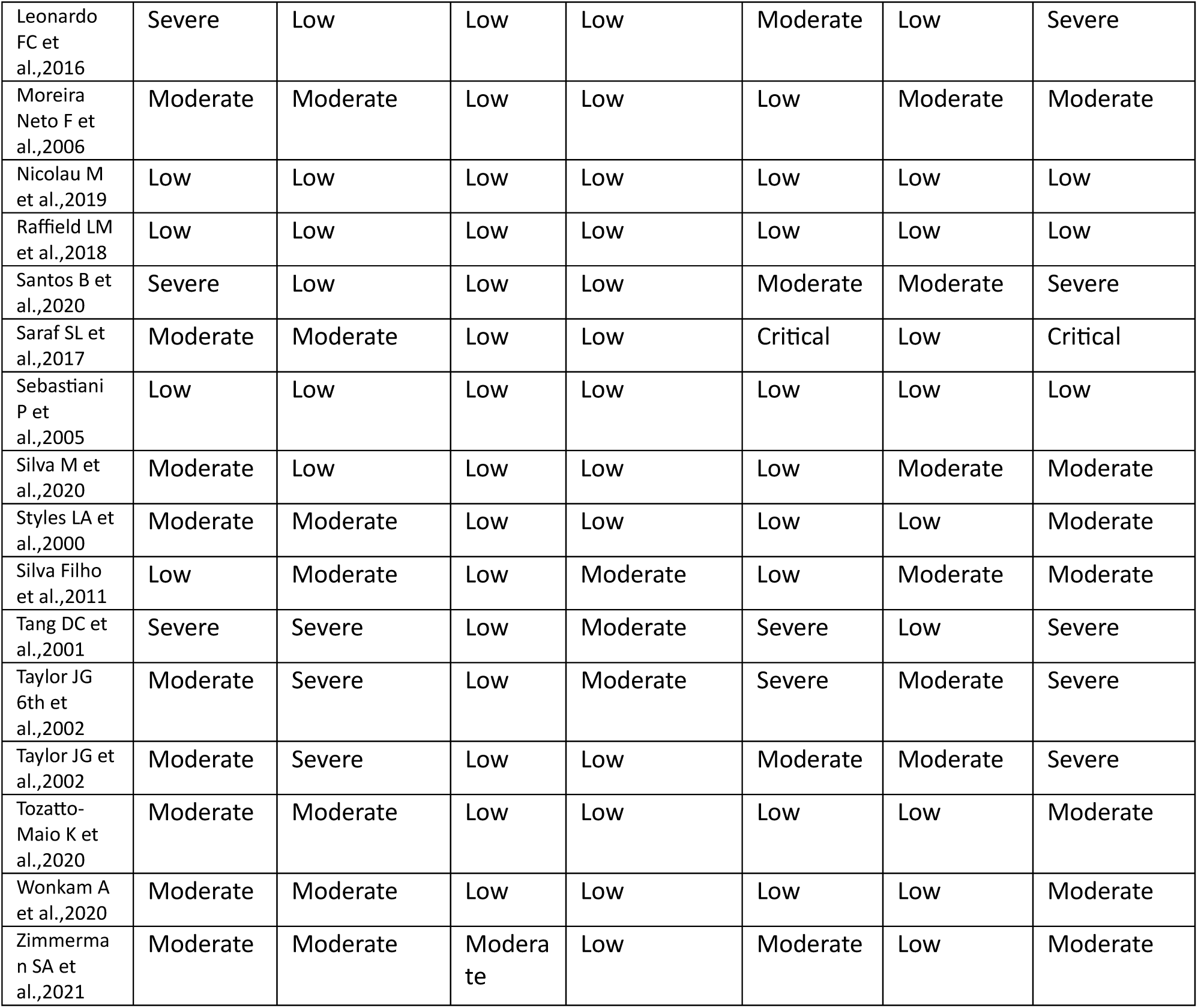
Risk of bias assessment for the studies on genetic factors associated with stroke in sickle cell disease.

## 4. Discussion

Here we identified 39 studies that performed an association of genetic variants with stroke in SCD patients, meta-analysis was possible for 12 loci of which genetic variants in *3.7kb α- thalassemia* deletion*, TEK*, *ADCY9,* and *ANXA2* were implicated to be significantly associated with stroke. Alpha-thalassemia and an SNP in the *ADCY9* gene protected against stroke while polymorphisms in TEK and ANXA2 genes were the risk factors for developing stroke in SCD.

Out of the 39 studies in this meta-analysis, 19 represented mixed ethnicities whereas 16 studies represented the population of African descent. Sub-Saharan Africa, the Mediterranean basin, the Middle East, and India are among the regions with the highest prevalence of SCD (4). Out of these regions, only individuals of African ancestry seem to be well represented in genetic studies that have explored the association for stroke among SCD patients. There was one study each which included a representative population from the Mediterranean basin and the Middle East but none from India. Studies from Brazil and USA represented the significant shares of the genetic studies on SCD patients where stroke was the outcome. As stroke represents one of the major complications associated with SCD, ethnic representation of major regions suffering from SCD in genetic studies of stroke in SCD will give a better picture of the loci implicated in this complication of SCD. The sample size of the majority of the studies evaluated here has been small; only a couple of studies have sample sizes crossing the 1000 mark. SCD might be one of the most common monogenic conditions but it is still a rare disease and hence such small numbers for genetic studies of a complication like stroke in this rare disorder is expected. However, the effect estimates of the genetic variants are still small for association with stroke in SCD, and hence collaboration of multiple studies and pooling of samples is required. Though there has been use of newer comprehensive genome-wide techniques like GWAS, whole exome sequencing, and whole genome sequencing studies for various complications in SCD patients for stroke there are only three whole exome/genome sequencing studies of small sample sizes. Unlike other rare disease conditions where collaborative efforts have led to the coming together of smaller studies to produce large meta-analyses, the same is still lacking in SCD, especially for complications including stroke.

α-Thalassemia in SCD patients is a protective factor against the risk of stroke (43,44). Multiple studies have implicated *3.7kb α-thalassemia deletion* that encompassed α- hemoglobin genes *HBA1* and *HBA2* as a protective factor against stroke in SCD. The meta-analysis of the six studies evaluated here also supports the protective role of the *α3.7kb deletion* (OR 0.46, 95% CI 0.34-0.61). We performed a random effect meta-analysis to find out whether the results of the fixed effect meta-analysis hold true. We observed that the association remained significant even after the Bonferroni correction.

*ADCY9* gene is located at 16p13.3 and encodes type 9 adenylate cyclase (AC). The enzyme is widely distributed in tissues and is critical for neuronal signalling(45). In our meta-analysis, we observed that *rs2238432-ADCY9* provided statistically significant protection from stroke (OR 0.45, 95% CI 0.29-0.72). This might be due to the anti-inflammatory cytokine production associated with the enzyme activity of type 9 AC.

Annexin A2 (ANXA2) gene located on chromosome 15q22.2 plays a key role in the coagulation processes(46) and is implicated as a genetic risk factor for stroke in SCD. In our random effect meta-analysis, we found that *rs11853426- ANXA2* (OR 1.97, 95% CI 1.25- 3.11) was significantly associated with the risk of stroke in sickle cell disease. Similarly, TEK gene is located at chromosome 9p21.2 and encodes TEK receptor tyrosine kinase. The protein facilitates communication between vascular endothelial cells and smooth muscle cells, ensures structural integrity of blood vessels and is involved in angiogenesis(47). We observed a high risk of developing stroke (OR 2.46, 95% CI 1.45-4.17) with rs489347-TEK genotype.

In a study by Flanagan JM et al, out of 38 SNPs that were investigated, five had a significant association with stroke in patients with sickle cell anemia. An SNP in the *ADCY9* gene and α- thalassemia decreased the risk of stroke in SCD whereas SNPs in the *TEK*, *ANXA2*, and *TGFBR3* genes increased stroke risk(23).

Although high fetal haemoglobin (HbF) is closely linked to a reduced rate of several vascular complications in SCD, it demonstrates a lack of protective effect on stroke in SCD(48). A mutation in the BCL11A gene that codes for BCL11A protein, a transcriptional repressor modulating the level of HbF, failed to show an association with painful episodes or other complications including stroke in SCD(49). Our meta-analysis observed that the genetic variation *rs1427407-BCL11A* has a weak protective effect against stroke in SCD (OR 0.89, 95% CI 0.57-1.04); however, it was statistically not significant. Similarly, variations in ENPP1, GOLGB1, VCAM1, PON1, MTHFR, TNF-α, and TGFBR3 genes also after Bonferroni correction, did not show a statistically significant association with stroke in SCD. The small sample size of the included studies might have contributed to a smaller effect size and lack of association of the SNP *rs284875-TGFBR3* with stroke in our meta-analysis.

Inclusion of larger studies is required to get a clearer picture of an association of -α3.7kb *Alpha-thalassemia deletion,* rs2238432-*ADCY*9, rs11853426-*ANXA2,* rs489347-*TEK* as well as other genotypes implicated in the causation of stroke in SCD patients. Larger genome-wide association studies may provide stronger evidence of the role of genetic variations in the causation of stroke in SCD. It is also of note that several susceptibility alleles likely act together to cause stroke in the study population. Therefore, risk prediction tools that integrate genetic risk factors, environmental risk factors, and other classical risk factors may provide important insights into the occurrence of stroke in SCD.

## 4.1 Strength and study limitations

The strength of our meta-analysis is that it is the first meta-analysis of studies exploring an association of genetic variants with stroke. Also, the lack of statistical heterogeneity among the included studies, the moderate overall methodological quality of the included studies, and a stringent adjustment of statistical significance test for multiple comparisons provide a shred of strong evidence for the association of the loci for risk prediction of stroke.

An important limitation of individual meta-analyses in this review includes a limited number of studies with small sample sizes in individual studies thereby limiting the ability to draw robust conclusions. Secondly, heterogeneity existed in the study population in terms of stroke definition and ethnicity. The majority of the studies were from mixed ethnicity. The present meta-analysis was restricted to articles published in the English language. As our literature survey identified 20 non-English studies, including 18 French, one German, and one Spanish article, it is likely that the interpretations of our meta-analyses are affected by the exclusion of those published articles.

## 5. Conclusion

The meta-analysis highlights the association of four genetic loci- -α3.7kb *Alpha-thalassemia deletion,* rs489347-*TEK*, rs2238432-*ADCY*9, and rs11853426-*ANXA2* with stroke in SCD.

## Data Availability

The raw data supporting the conclusions of this research work is presented in the article or in the supplementary material provided with the article.

## Acknowledgment

We acknowledge our institute, Rajendra Institute of Medical Sciences, Ranchi for providing the resources for conducting this study.

## Abbreviations and acronyms

SCD: sickle cell disease
OR: odds ratio
CI: confidence interval
HbF: fetal hemoglobin
GWAS: genome-wide association studies
WES: Whole exome sequencing
SNP: single nucleotide polymorphism
AC: adenylyl cyclase
WHO: World Health Organization

## References

1. Platt OS, Brambilla DJ, Rosse WF, Milner PF, Castro O, Steinberg MH, et al. Mortality In Sickle Cell Disease -- Life Expectancy and Risk Factors for Early Death. N Engl J Med. 1994 Jun 9;330(23):1639–44.

2. Piel FB, Hay SI, Gupta S, Weatherall DJ, Williams TN. Global Burden of Sickle Cell Anaemia in Children under Five, 2010–2050: Modelling Based on Demographics, Excess Mortality, and Interventions. Osrin D, editor. PLoS Med. 2013 Jul 16;10(7):e1001484.

3. Sedrak A, Kondamudi NP. Sickle Cell Disease. In: StatPearls [Internet]. Treasure Island (FL): StatPearls Publishing; 2023 [cited 2023 Aug 8]. Available from: http://www.ncbi.nlm.nih.gov/books/NBK482384/

4. Cerebrovascular Accidents in Sickle Cell Disease: Rates and Risk Factors - ScienceDirect [Internet]. [cited 2022 May 28]. Available from: https://www.sciencedirect.com/science/article/pii/S0006497120549082?via%3Dihub

5. Piel FB, Steinberg MH, Rees DC. Sickle Cell Disease. N Engl J Med. 2017 Apr 20;376(16):1561–73.

6. de Melo TRF, Ercolin L dos R, Chelucci RC, Melchior ACB, Lanaro C, Chin CM, et al. Sickle Cell Disease – Current Treatment and New Therapeutical Approaches. In: Munshi A, editor. Inherited Hemoglobin Disorders [Internet]. InTech; 2015 [cited 2022 May 28]. Available from: http://www.intechopen.com/books/inherited-hemoglobin-disorders/sickle-cell-disease-current-treatment-and-new-therapeutical-approaches

7. Nicolau M, Vargas S, Silva M, Coelho A, Ferreira E, Mendonça J, et al. Genetic modulators of fetal hemoglobin expression and ischemic stroke occurrence in African descendant children with sickle cell anemia. Ann Hematol. 2019 Dec;98(12):2673–81.

8. Silva M, Vargas S, Coelho A, Ferreira E, Mendonça J, Vieira L, et al. Biomarkers and genetic modulators of cerebral vasculopathy in sub-Saharan ancestry children with sickle cell anemia. Blood Cells Mol Dis. 2020 Jul;83:102436.

9. Belisário AR, Nogueira FL, Rodrigues RS, Toledo NE, Cattabriga ALM, Velloso-Rodrigues C, et al. Association of alpha-thalassemia, TNF-alpha (-308G>A) and VCAM-1 (c.1238G>C) gene polymorphisms with cerebrovascular disease in a newborn cohort of 411 children with sickle cell anemia. Blood Cells Mol Dis. 2015 Jan;54(1):44–50.

10. Franco RS, Yasin Z, Palascak MB, Ciraolo P, Joiner CH, Rucknagel DL. The effect of fetal hemoglobin on the survival characteristics of sickle cells. Blood. 2006 Aug 1;108(3):1073–6.

11. Adekile A. The Genetic and Clinical Significance of Fetal Hemoglobin Expression in Sickle Cell Disease. Med Princ Pract. 2021;30(3):201–11.

12. Sidani CA, Ballourah W, El Dassouki M, Muwakkit S, Dabbous I, Dahoui H, et al. Venous sinus thrombosis leading to stroke in a patient with sickle cell disease on hydroxyurea and high hemoglobin levels: Treatment with thrombolysis. Am J Hematol. 2008 Oct;83(10):818–20.

13. Heit JA, Armasu SM, McCauley BM, Kullo IJ, Sicotte H, Pathak J, et al. Identification of unique venous thromboembolism-susceptibility variants in African-Americans. Thromb Haemost. 2017 Apr 3;117(4):758–68.

14. Hoppe C, Klitz W, D’Harlingue K, Cheng S, Grow M, Steiner L, et al. Confirmation of an Association Between the TNF(−308) Promoter Polymorphism and Stroke Risk in Children With Sickle Cell Anemia. Stroke. 2007 Aug;38(8):2241–6.

15. Flanagan JM, Sheehan V, Linder H, Howard TA, Wang YD, Hoppe CC, et al. Genetic mapping and exome sequencing identify 2 mutations associated with stroke protection in pediatric patients with sickle cell anemia. Blood. 2013 Apr 18;121(16):3237–45.

16. Hoppe C, Cheng S, Grow M, Silbergleit A, Klitz W, Trachtenberg E, et al. A novel multilocus genotyping assay to identify genetic predictors of stroke in sickle cell anaemia: Short Report. British Journal of Haematology. 2001 Sep;114(3):718–20.

17. Belisário AR, Sales RR, Toledo NE, Velloso-Rodrigues C, Silva CM, Viana MB. Association between ENPP1 K173Q and stroke in a newborn cohort of 395 Brazilian children with sickle cell anemia. Blood. 2015 Sep 3;126(10):1259–60.

18. Belisário AR, Sales RR, Toledo NE, Muniz MB de SR, Velloso-Rodrigues C, Silva CM, et al. Reticulocyte count is the most important predictor of acute cerebral ischemia and high-risk transcranial Doppler in a newborn cohort of 395 children with sickle cell anemia. Ann Hematol. 2016 Oct;95(11):1869–80.

19. Cumming AM, Olujohungbe A, Keeney S, Singh H, Hay CR, Serjeant GR. The methylenetetrahydrofolate reductase gene C677T polymorphism in patients with homozygous sickle cell disease and stroke. Br J Haematol. 1999 Dec;107(3):569–71.

20. Neto F, Enzo LDDML, Noguti MAE, Morelli VM, Gil ICP, Beltrão ACS, et al. The clinical impact of MTHFR polymorphism on the vascular complications of sickle cell disease. Brazilian Journal of Medical and Biological Research. 2006 Oct 1;39.

21. Hatzlhofer BLD, Pereira-Martins DA, de Farias Domingos I, Arcanjo G da S, Weinhäuser I, Falcão DA, et al. Alpha thalassemia, but not βS-globin haplotypes, influence sickle cell anemia clinical outcome in a large, single-center Brazilian cohort. Ann Hematol. 2021 Apr;100(4):921–31.

22. Zimmerman SA, Howard TA, Whorton MR, Rosse WF, James AH, Ware RE. Thrombophilic DNA Mutations As Independent Risk Factors for Stroke and Avascular Necrosis in Sickle Cell Anemia. Hematology. 2001;6(5):347–53.

23. Vi JGT, Tang DC, Savage SA, Leitman SF, Heller SI, Serjeant GR, et al. Variants in the VCAM1 gene and risk for symptomatic stroke in sickle cell disease. Blood. 2002 Dec 15;100(13):4303–9.

24. Flanagan JM, Frohlich DM, Howard TA, Schultz WH, Driscoll C, Nagasubramanian R, et al. Genetic predictors for stroke in children with sickle cell anemia. Blood. 2011 Jun 16;117(24):6681–4.

25. Batista JVGF, Pereira-Martins DA, Falcão DA, Domingos IF, Arcanjo GS, Hatzlhofer BL, et al. Association of KLOTHO polymorphisms with clinical complications of sickle cell anemia. Ann Hematol. 2021 Aug;100(8):1921–7.

26. Domingos IF, Falcão DA, Hatzlhofer BL, Cunha AF, Santos MN, Albuquerque DM, et al. Influence of the βs haplotype and α-thalassemia on stroke development in a Brazilian population with sickle cell anaemia. Ann Hematol. 2014 Jul;93(7):1123–9.

27. Domingos IF, Pereira-Martins DA, Borges-Medeiros RL, Falcao DA, Hatzlhofer BL, Brewin JN, et al. Evaluation of oxidative stress-related genetic variants for predicting stroke in patients with sickle cell anemia. J Neurol Sci. 2020 Jul 15;414:116839.

28. Hassan FM, Al-Zahrani FM. BCL11A rs1427407 Genotypes in Sickle Cell Anemia Patients Undergo to Stroke Problems in Sudan. Korean J Fam Med. 2019 Jan;40(1):53–7.

29. Bhatnagar P, Barron-Casella E, Bean CJ, Milton JN, Baldwin CT, Steinberg MH, et al. Genome-wide meta-analysis of systolic blood pressure in children with sickle cell disease. PLoS One. 2013;8(9):e74193.

30. Brewin JN, Rooks H, Gardner K, Senior H, Morje M, Patel H, et al. Genome wide association study of silent cerebral infarction in sickle cell disease (HbSS and HbSC). Haematologica. 2021 Jun 1;106(6):1770–3.

31. Alsultan A, Alabdulaali MK, Griffin PJ, Alsuliman AM, Ghabbour HA, Sebastiani P, et al. Sickle cell disease in Saudi Arabia: the phenotype in adults with the Arab-Indian haplotype is not benign. Br J Haematol. 2014 Feb;164(4):597–604.

32. Raffield LM, Zakai NA, Duan Q, Laurie C, Smith JD, Irvin MR, et al. D-Dimer in African Americans: Whole Genome Sequence Analysis and Relationship to Cardiovascular Disease Risk in the Jackson Heart Study. Arterioscler Thromb Vasc Biol. 2017 Nov;37(11):2220–7.

33. Coupland AP, Thapar A, Qureshi MI, Jenkins H, Davies AH. The definition of stroke. J R Soc Med. 2017 Jan;110(1):9–12.

34. Willer CJ, Li Y, Abecasis GR. METAL: fast and efficient meta-analysis of genomewide association scans. Bioinformatics. 2010 Sep 1;26(17):2190–1.

35. Assessing the quality of published genetic association studies in meta-analyses: the quality of genetic studies (Q-Genie) tool | BMC Genomic Data | Full Text [Internet]. [cited 2023 May 10]. Available from: https://bmcgenomdata.biomedcentral.com/articles/10.1186/s12863-015-0211-2

36. Sterne JA, Hernán MA, Reeves BC, Savović J, Berkman ND, Viswanathan M, et al. ROBINS-I: a tool for assessing risk of bias in non-randomised studies of interventions. BMJ. 2016 Oct 12;355:i4919.

37. Ryan-Moore E, Mavrommatis Y, Waldron M. Systematic Review and Meta-Analysis of Candidate Gene Association Studies With Fracture Risk in Physically Active Participants. Front Genet. 2020 Jun 16;11:551.

38. Raffield LM, Ulirsch JC, Naik RP, Lessard S, Handsaker RE, Jain D, et al. Common α-globin variants modify hematologic and other clinical phenotypes in sickle cell trait and disease. PLoS Genet. 2018 Mar;14(3):e1007293.

39. Sebastiani P, Ramoni MF, Nolan V, Baldwin CT, Steinberg MH. Genetic dissection and prognostic modeling of overt stroke in sickle cell anemia. Nat Genet. 2005 Apr;37(4):435–40.

40. Hatzlhofer BLD, Bezerra MAC, Santos MNN, Albuquerque DM, Freitas EM, Costa FF, et al. MTHFR polymorphic variant C677T is associated to vascular complications in sickle-cell disease. Genet Test Mol Biomarkers. 2012 Sep 1;16(9):1038–43.

41. Tang DC, Prauner R, Liu W, Kim KH, Hirsch RP, Driscoll MC, et al. Polymorphisms within the angiotensinogen gene (GT-repeat) and the risk of stroke in pediatric patients with sickle cell disease: a case-control study. Am J Hematol. 2001 Nov;68(3):164–9.

42. Taylor Vi JG, Tang D, Foster CB, Serjeant GR, Rodgers GP, Chanock SJ. Patterns of low-affinity immunoglobulin receptor polymorphisms in stroke and homozygous sickle cell disease. Am J Hematol. 2002 Feb;69(2):109–14.

43. Santos B, Delgadinho M, Ferreira J, Germano I, Miranda A, Arez AP, et al. Co-Inheritance of alpha-thalassemia and sickle cell disease in a cohort of Angolan pediatric patients. Mol Biol Rep. 2020 Jul;47(7):5397–402.

44. Ali Al-Barazanchi ZA, Abdulateef SS, Hassan MK. Co-Inheritance of α-thalassemia gene mutation in patients with sickle cell Disease: Impact on clinical and hematological variables. Niger J Clin Pract. 2021 Jun;24(6):874–82.

45. Rhainds D, Packard CJ, Brodeur MR, Niesor EJ, Sacks FM, Jukema JW, et al. Role of Adenylate Cyclase 9 in the Pharmacogenomic Response to Dalcetrapib: Clinical Paradigm and Molecular Mechanisms in Precision Cardiovascular Medicine. Circ Genom Precis Med. 2021 Apr;14(2):e003219.

46. The annexin A2 system and angiogenesis - PubMed [Internet]. [cited 2023 Aug 2]. Available from: https://pubmed.ncbi.nlm.nih.gov/27366903/

47. TEK TEK receptor tyrosine kinase - NIH Genetic Testing Registry (GTR) - NCBI [Internet]. [cited 2023 Aug 2]. Available from: https://www.ncbi.nlm.nih.gov/gtr/genes/7010/

48. Steinberg MH, Sebastiani P. Genetic Modifiers of Sickle Cell Disease. Am J Hematol. 2012 Aug;87(8):795–803.

49. Chang AK, Ginter Summarell CC, Birdie PT, Sheehan VA. Genetic modifiers of severity in sickle cell disease. Clin Hemorheol Microcirc. 2018;68(2–3):147–64.

50. Alsultan A, Al-Suliman AM, Aleem A, AlGahtani FH, Alfadhel M. Utilizing Whole-Exome Sequencing to Characterize the Phenotypic Variability of Sickle Cell Disease. Genet Test Mol Biomarkers. 2018 Sep;22(9):561–7.

51. Andrade FL, Annichino-Bizzacchi JM, Saad ST, Costa FF, Arruda VR. Prothrombin mutant, factor V Leiden, and thermolabile variant of methylenetetrahydrofolate reductase among patients with sickle cell disease in Brazil. Am J Hematol. 1998 Sep;59(1):46–50.

52. Barbosa LC, Miranda-Vilela AL, Hiragi C de O, Ribeiro IF, Daldegan MB, Grisolia CK, et al. Haptoglobin and myeloperoxidase (- G463A) gene polymorphisms in Brazilian sickle cell patients with and without secondary iron overload. Blood Cells Mol Dis. 2014 Mar;52(2–3):95–107.

53. Brewin JN, Smith AE, Cook R, Tewari S, Brent J, Wilkinson S, et al. Genetic Analysis of Patients With Sickle Cell Anemia and Stroke Before 4 Years of Age Suggest an Important Role for Apoliprotein E. Circ Genom Precis Med. 2020 Oct;13(5):531–40.

54. de Oliveira Filho RA, Silva GJ, de Farias Domingos I, Hatzlhofer BLD, da Silva Araújo A, de Lima Filho JL, et al. Association between the genetic polymorphisms of glutathione S-transferase (GSTM1 and GSTT1) and the clinical manifestations in sickle cell anemia. Blood Cells, Molecules, and Diseases. 2013 Aug 1;51(2):76–9.

55. Domingos IF, Pereira-Martins DA, Coelho-Silva JL, Borges-Medeiros RL, Falcão DA, Azevedo RC, et al. Interleukin-6 G-174C polymorphism predicts higher risk of stroke in sickle cell anaemia. Br J Haematol. 2018 Jul;182(2):294–7.

56. ElAlfy MS, Ebeid FSE, Kamal TM, Eissa DS, Ismail EAR, Mohamed SH. Angiotensinogen M235T Gene Polymorphism is a Genetic Determinant of Cerebrovascular and Cardiopulmonary Morbidity in Adolescents with Sickle Cell Disease. J Stroke Cerebrovasc Dis. 2019 Feb;28(2):441–9.

57. Hoppe C, Klitz W, Noble J, Vigil L, Vichinsky E, Styles L. Distinct HLA associations by stroke subtype in children with sickle cell anemia. Blood. 2003 Apr 1;101(7):2865–9.

58. Hoppe C, Klitz W, Cheng S, Apple R, Steiner L, Robles L, et al. Gene interactions and stroke risk in children with sickle cell anemia. Blood. 2004 Mar 15;103(6):2391–6.

59. Leonardo FC, Brugnerotto AF, Domingos IF, Fertrin KY, de Albuquerque DM, Bezerra MAC, et al. Reduced rate of sickle-related complications in Brazilian patients carrying HbF-promoting alleles at the BCL11A and HMIP-2 loci. Br J Haematol. 2016 May;173(3):456–60.

60. Saraf SL, Akingbola TS, Shah BN, Ezekekwu CA, Sonubi O, Zhang X, et al. Associations of α- thalassemia and BCL11A with stroke in Nigerian, United States, and United Kingdom sickle cell anemia cohorts. Blood Adv. 2017 Apr 25;1(11):693–8.

61. Filho ILDS, Leite ACCB, Moura PG, Ribeiro GS, Cavalcante AC, Azevedo FCMD, et al. Genetic polymorphisms and cerebrovascular disease in children with sickle cell anemia from Rio de Janeiro, Brazil. Arq Neuro-Psiquiatr. 2011 Jun;69(3):431–5.

62. Styles LA, Hoppe C, Klitz W, Vichinsky E, Lubin B, Trachtenberg E. Evidence for HLA-related susceptibility for stroke in children with sickle cell disease. Blood. 2000 Jun 1;95(11):3562–7.

63. Tozatto-Maio K, Girot R, Ly ID, Silva Pinto AC, Rocha V, Fernandes F, et al. Polymorphisms in Inflammatory Genes Modulate Clinical Complications in Patients With Sickle Cell Disease. Front Immunol. 2020;11:2041.

64. Wonkam A, Chimusa ER, Mnika K, Pule GD, Ngo Bitoungui VJ, Mulder N, et al. Genetic modifiers of long-term survival in sickle cell anemia. Clin Transl Med. 2020 Aug;10(4):e152.

65. Genetics against race: Science, politics and affirmative action in Brazil - PubMed [Internet]. [cited 2023 Aug 10]. Available from: https://pubmed.ncbi.nlm.nih.gov/27479998/

